# Incident psoriasis in atopic dermatitis: A large-scale cohort study of disease- and treatment-associated risks

**DOI:** 10.64898/2026.04.18.26351181

**Authors:** Florian Thaqi, Katja Bieber, Hatim Kerniss, Khalaf Kridin, Philip Curman, Ralf J. Ludwig

**Author notes:** equal contributors (Philip Curman, Ralf J. Ludwig). **Declaration of interest**: PC has received travel grants from TriNetX. RJL has received travel grants from TriNetX and has received research funding from Euroimmun, Dompe pharma, Novartis and Sanofi. All other authors declare no relevant conflict of interest.

## Abstract

**Background:** Clinical and genetic evidence on the association between atopic dermatitis (AD) and subsequent psoriasis remains conflicting, and it is unclear whether this risk is modified by systemic treatments. Recent reports suggest type 2-targeted biologics may unmask psoriasis in AD patients, but data are limited. We thus aimed to assess whether AD is associated with incident psoriasis and whether this risk differs by systemic treatment, particularly biologics versus conventional systemic immunosuppressants (cvIS).

**Methods:** Scoping analyses informed a locked analytic design, preregistration at OSF, and confirmatory execution. Propensity score–matched analyses compared AD with non-AD controls and biologics with cvIS. Sensitivity analyses, Cox model triangulation, and control outcomes assessed robustness.

**Findings:** Among ∼300,000 matched pairs, AD was associated with increased psoriasis risk (primary HR 3.81, 95% CI 3.35-4.34), consistent across all 8 sensitivity analyses and model triangulation. Biologic treatment was associated with reduced psoriasis risk versus cvIS (primary HR 0.20, 95% CI 0.11–0.35), consistent across 6 of 7 evaluable sensitivity analyses and Cox triangulation. Positive and negative control outcomes showed expected directional patterns.

**Interpretation:** Acknowledging limitations including residual confounding and coding misclassification, AD was associated with increased psoriasis risk and biologics with lower psoriasis risk than cvIS.

**Funding:** DFG (EXC2167, SFB1526, LU877/25-1), Schleswig-Holstein Excellence-Chair Program, Swedish Society for Dermatology and Venereology, and the Tore Nilson Foundation.

**Research in context:** *Evidence before this study:* Atopic dermatitis (eczema) and psoriasis are the two most common chronic inflammatory skin diseases worldwide. For a long time, doctors and researchers assumed these two conditions could not occur in the same person, as they were thought to involve opposing immune responses. However, this view has been challenged over the past decade. Some large studies, including population-based cohorts from Taiwan and the United Kingdom, have found that people with eczema may be at higher risk of developing psoriasis over time, while other studies, including genetic analyses, have suggested the opposite: that the two diseases may actually protect against each other. This conflicting picture has left clinicians uncertain about the true relationship between the two diseases in everyday clinical practice. A separate but related concern has emerged with the introduction of a new class of highly effective treatments for eczema, biologics, particularly dupilumab. Case reports and observational studies, including a large study published in JAMA Dermatology in 2025, have raised the possibility that these medications might trigger psoriasis in some patients, potentially by shifting the immune system from one inflammatory pattern to another. However, prior studies on this question had important methodological limitations: they were not pre-planned and registered before data collection, they did not always tightly link treatment use to an eczema diagnosis, and critically, none compared biologic treatment directly against conventional immunosuppressant medications, the most relevant clinical comparator.

*Added value of this study:* This study is a large and methodologically rigorous investigation of both questions: whether eczema itself increases the risk of developing psoriasis, and whether the type of systemic treatment used for eczema influences that risk. Using a database of over 110 million electronic health records from across the United States, we matched approximately 300,000 patients with eczema to 300,000 patients without eczema and followed them for up to seven years. We also compared nearly 5,500 patients treated with biologics to an equal number treated with conventional immunosuppressants. Crucially, our study was pre-registered before any data were analyzed, meaning the research questions, methods, and analyses were locked in advance and could not be adjusted based on what the data showed. We also used a range of additional analyses to test whether our findings were robust, including checks using outcomes that should not be affected by eczema or its treatment (such as appendectomy and hearing loss), which confirmed that our results were not likely explained by bias alone. We found that eczema was associated with an increased risk of developing psoriasis, but that this risk was substantially influenced by the choice of comparison group, ranging from approximately 1.4-fold to nearly 4-fold depending on the analytical approach. More strikingly, we found that patients treated with biologics had a markedly lower risk of developing psoriasis compared with those treated with conventional immunosuppressants, the opposite of what prior reports had suggested. This finding was consistent across nearly all additional analyses performed.

*Implications of all the available evidence:* Taken together with existing evidence, these findings suggest two important conclusions. First, clinicians should be aware that eczema, particularly moderate-to-severe eczema requiring systemic treatment, may carry an elevated risk of developing psoriasis over time. This does not mean that all patients with eczema need to be screened for psoriasis routinely, but it does support clinical awareness and monitoring in higher-risk patients. Second, and perhaps most importantly for treatment decisions, biologics do not appear to increase the risk of psoriasis compared with conventional immunosuppressants and may in fact be associated with a lower risk. This provides reassurance for patients and clinicians considering biologic therapy and challenges the narrative that these medications trigger psoriasis. Future research should aim to confirm these findings in other populations, investigate the biological mechanisms underlying the relationship between eczema and psoriasis, and examine whether specific biologic agents differ from one another in their effects on psoriasis risk.

## Introduction

Atopic dermatitis (AD) and psoriasis are the two most prevalent chronic inflammatory skin diseases (CISD), together affecting over 10% of the global population. Traditionally, they have been considered immunologically distinct: AD being characterized by type 2 helper T (Th2) cell-mediated inflammation with elevated IL-4 and IL-13, whereas psoriasis is driven by Th1/Th17 responses with predominant IL-17 and IL-22 production (1,2). In support of this notion, a key study demonstrated that patients with concurrent AD and psoriasis exhibited distinct T-cell infiltrates in lesions from each disease, with an antagonistic clinical course (3). However, this traditional dichotomy has been challenged by observations of phenotypic overlap, mixed immunophenotypes, and disease transition (4–6).

Epidemiological evidence on the relationship between AD and subsequent psoriasis risk remains conflicting. Population-based cohort studies from Taiwan have reported bidirectional associations, suggesting that patients with either disease may be at elevated risk for developing the other (7). A UK primary care cohort of over 170,000 AD patients similarly found increased risk of incident psoriatic arthritis, particularly in severe AD (8). In contrast, a German cross-sectional study of 90,000 dermatologist-examined individuals found type-2 inflammatory diseases, including AD, were significantly less frequent in persons with psoriasis, supporting the traditional mutual exclusivity hypothesis (9). Genetic evidence yields conflicting results. Mendelian randomization studies documented that AD did not affect psoriasis risk (10) – or that AD may act as a protective factor for psoriasis (11). Moreover, genetic analyses demonstrated that AD and psoriasis harbor opposing risk alleles at shared loci, with no evidence of shared loci acting in the same direction, supporting the notion of partial mutual exclusivity between the two diseases (12). This discordance in observational and genetic evidence leaves uncertainty regarding whether AD itself confers psoriasis risk in routine clinical care.

The emergence of type 2-targeted biologics, specifically dupilumab, tralokinumab, and lebrikizumab, has transformed AD management. However, case reports and pharmacovigilance signals have suggested that these biologics, especially dupilumab, may unmask or trigger psoriasis (termed as switch), potentially through Th2-to-Th17 immune skewing (13–15). In an Italian multicenter retrospective cohort study including 5,899 AD patients, psoriasis developed in 1.3% receiving dupilumab and 2.1% on tralokinumab (14). However, this study lacked a cvIS comparator, precluding direct assessment of whether psoriasis risk differs by treatment class. In line, a recent TriNetX cohort study reported a 58% higher psoriasis risk in adult AD treated with dupilumab as opposed to other systemic agents (16). However, methodological limitations warrant caution: Treatment exposure was not tightly anchored to AD diagnosis (potentially including patients treated for other indications), the analysis was restricted to adults, limiting generalizability to pediatric populations where AD is most prevalent, and some cohort definitions included immortal time bias. Furthermore, pre-registration was not reported, limiting assessment of whether analyses were hypothesis-driven versus exploratory (17). As such, comparative data evaluating biologics versus conventional systemic immunosuppressants (cvIS) with rigorous design features remain limited.

Given these knowledge gaps, we conducted a large-scale, pre-registered retrospective cohort study to address two aims: (1) to determine whether AD is associated with increased incident psoriasis risk compared with matched non-AD controls, and (2) to assess whether psoriasis risk among patients with AD differs according to systemic treatment exposure, particularly type 2-targeted biologics versus cvIS. This pre-registered study employed propensity score matching, pre-specified sensitivity analyses, multivariable Cox model triangulation, and positive and negative control outcomes to strengthen the robustness of observed associations.

## Methods

A detailed description of the methos used in provided in the Supplement Materials and Methods.

### Study design and data source

A propensity score–matched retrospective cohort study was conducted using the US Collaborative Network of the TriNetX federated EHR platform, following established procedures (18). The study comprised an exploratory scoping phase, design freeze, preregistration (OSF: https://osf.io/8f2s4/registrations, March 26, 2026), and confirmatory execution (March 27-28, 2026). At analysis, the network included over 110 million EHRs from 65–66 healthcare organizations. Reporting followed the STROBE guidelines (19). The analysis used de-identified data; institutional review board approval was not required.

### Preliminary scoping analysis and design finalization

A scoping analysis confirmed feasibility of cohort definitions and outcome ascertainment without informing confirmatory inference. For Aim 2, only biologic versus cvIS comparisons met the predefined ≥1,000 patient threshold; JAK inhibitor and IL-13 inhibitor cohorts were excluded. Following scoping, the analytic framework was locked (design freeze: March 26, 2026), preregistered, and executed without further modification.

### Study population and index events

<u>Aim 1</u>: Cases were patients with first recorded AD diagnosis (ICD-10-CM L20) between January 2016 and December 2018; controls had first general health examination (Z00) during the same period. Patients with prior psoriasis or control outcomes were excluded (Supplement Table 1). <u>Aim 2</u>. Treatment cohorts were defined from first systemic treatment on or after January 2017. The biologic cohort received dupilumab, tralokinumab, or lebrikizumab; the cvIS cohort received methotrexate, azathioprine, mycophenolate mofetil, or ciclosporin. AD diagnosis (L20) within 1 month before treatment was required. Prior exposure to reciprocal treatments or JAK inhibitors was excluded. Exposure was defined at first treatment record with follow-up irrespective of treatment changes (Supplement Table 2). Based on prespecified thresholds (≥1,000 patients), JAK inhibitor (n=983) and IL-13 inhibitor (n=430) comparisons were not performed.

### Outcomes

The primary outcome was incident psoriasis vulgaris (L40.0). Negative control outcomes included burns/corrosions, presbycusis, and appendectomy; topical betamethasone served as positive control in Aim 1. Additional preregistered outcomes (atopic dermatitis, acne, rosacea, hidradenitis suppurativa, vitiligo, alopecia areata, lichen planus, depression) were inadvertently carried over and not analyzed.

### Covariates and propensity score matching

Covariates selected a priori were included in propensity score models (Supplement Tables 1-2). Cohorts were matched 1:1 using greedy nearest-neighbor matching (caliper: 0.1 SD). Follow-up was fixed at 2,595 days (Aim 1) and 601 days (Aim 2) based on scoping.

### Sensitivity Analyses and Model Triangulation

Prespecified sensitivity analyses (eight for Aim 1, nine for Aim 2) assessed robustness, including lag-time analyses, stricter AD definitions, age stratification, and alternative comparators. model For model triangulation Multivariable Cox was performed. For Aim 1, computational constraints required substituting acne vulgaris controls (post-hoc modification).

### Statistical analysis

Baseline characteristics were compared using t-tests (continuous) and z-tests (categorical); balance was assessed using standardized mean differences. Missing covariate values were retained in all analyses, treated as a separate category in propensity score matching and retained in Cox models. Time-to-event analyses used Kaplan-Meier methods with log-rank tests. Bonferroni correction was applied (Aim 1: α=0.01; Aim 2: α=0.025).

### Role of the funding source

The funders had no role in study design, data collection, analysis, or interpretation, in the writing of the report, or in the decision to submit the paper for publication.

## Results

The results are presented in two parts: Aim 1 examines the association between atopic dermatitis and subsequent psoriasis risk, whereas Aim 2 evaluates psoriasis risk according to systemic treatment exposure in atopic dermatitis.

### Risk of subsequent psoriasis in patients with AD

In the primary analysis, 351,070 patients with AD and 6,888,769 non-AD controls were identified before matching. Propensity score matching yielded 351,069 matched pairs. Before matching, cohorts differed substantially; after matching, all baseline covariates were well balanced (SMDs <0.10). Median follow-up was 2,595 days in both cohorts (interquartile range: 1,219 [AD] and 1,129 days [controls]). Full baseline characteristics are presented in Supplement Table 3.

Psoriasis was documented in 1,088 of 351,069 patients with AD (0.31%) versus 290 of 351,069 matched controls (0.08%). Kaplan–Meier analysis demonstrated higher cumulative incidence in the AD cohort (p<0.0001, HR 3.81, 95%-CI, 3.35-4.34, Figure 1, Supplement Table 3). AD was associated with an increased risk of subsequent psoriasis. Comparator choice influenced the magnitude of this association. In sensitivity analyses using Z00 controls, HRs ranged from 3.07 to 4.04, whereas substituting acne vulgaris as an active comparator yielded a more conservative estimate (HR 1.73, 95%-CI, 1.56-1.93), and Cox model triangulation using the acne comparator similarly attenuated the association (HR 1.39, 95%-CI, 1.27-1.52, p<0.001, Table 1). This attenuation likely reflects partial adjustment for healthcare-seeking behavior and dermatology-specific coding patterns shared between AD and acne patients. Notwithstanding this comparator-dependent variation, the direction of association was consistent across all prespecified sensitivity analyses (HR range: 1.39–4.04, Supplement Table 3). The association also persisted after applying a 3-month lag time (HR 3.61, 95%-CI, 3.16-4.12), restricting to patients with at least 6 months of baseline data (HR 3.48, 95%-CI, 3.03-3.99), using stricter AD definitions with and without potential immortal time bias (HR 4.04, 95%-CI, 3.20-5.12 and HR 3.41, 95%-CI, 3.01-3.86, respectively), and excluding patients with pre-existing inflammatory skin diseases (HR 3.07, 95%-CI, 2.53-3.73). The association was observed in both age strata: patients younger than 18 years (HR 2.86, 95%-CI, 2.28-3.59) and those aged 18 years or older (HR 3.41, 95%-CI, 2.95-3.94).

**Figure 1.**
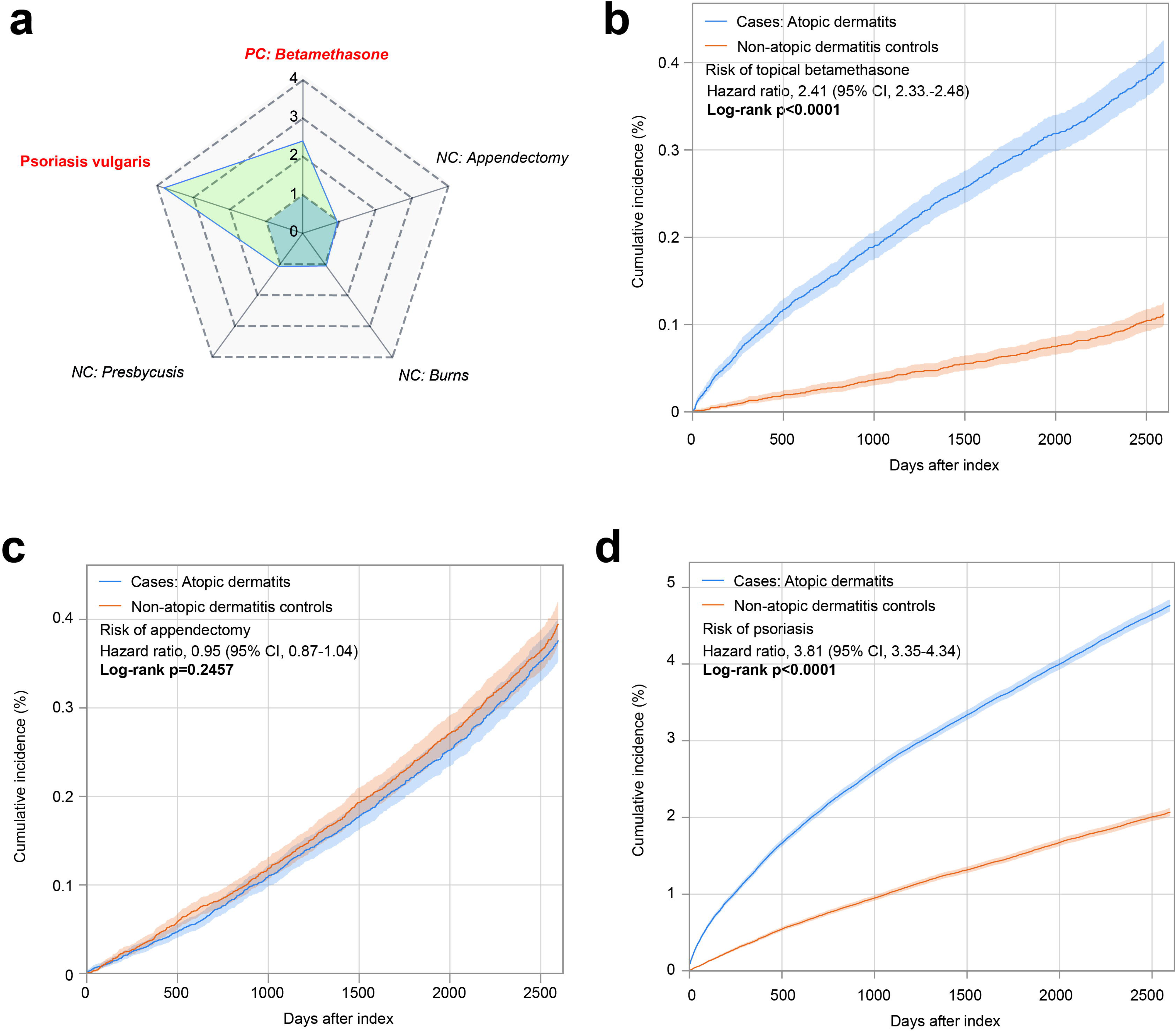
Primary outcome and control outcome analyses in patients with atopic dermatitis versus matched non-atopic dermatitis controls. (**a**) Radar plot depicting hazard ratios (HRs) from the primary analysis across all evaluated endpoints. Positive control (PC) and negative control (NC) outcomes are indicated in italics; red labels denote statistically significant associations. (**b-d**) Kaplan-Meier curves showing cumulative incidence of (**b**) topical betamethasone exposure (positive control), (**c**) appendectomy (negative control), and (**d**) psoriasis vulgaris (primary outcome) in patients with AD and matched non-AD controls. Shaded bands represent 95% confidence intervals.

**Table 1.**
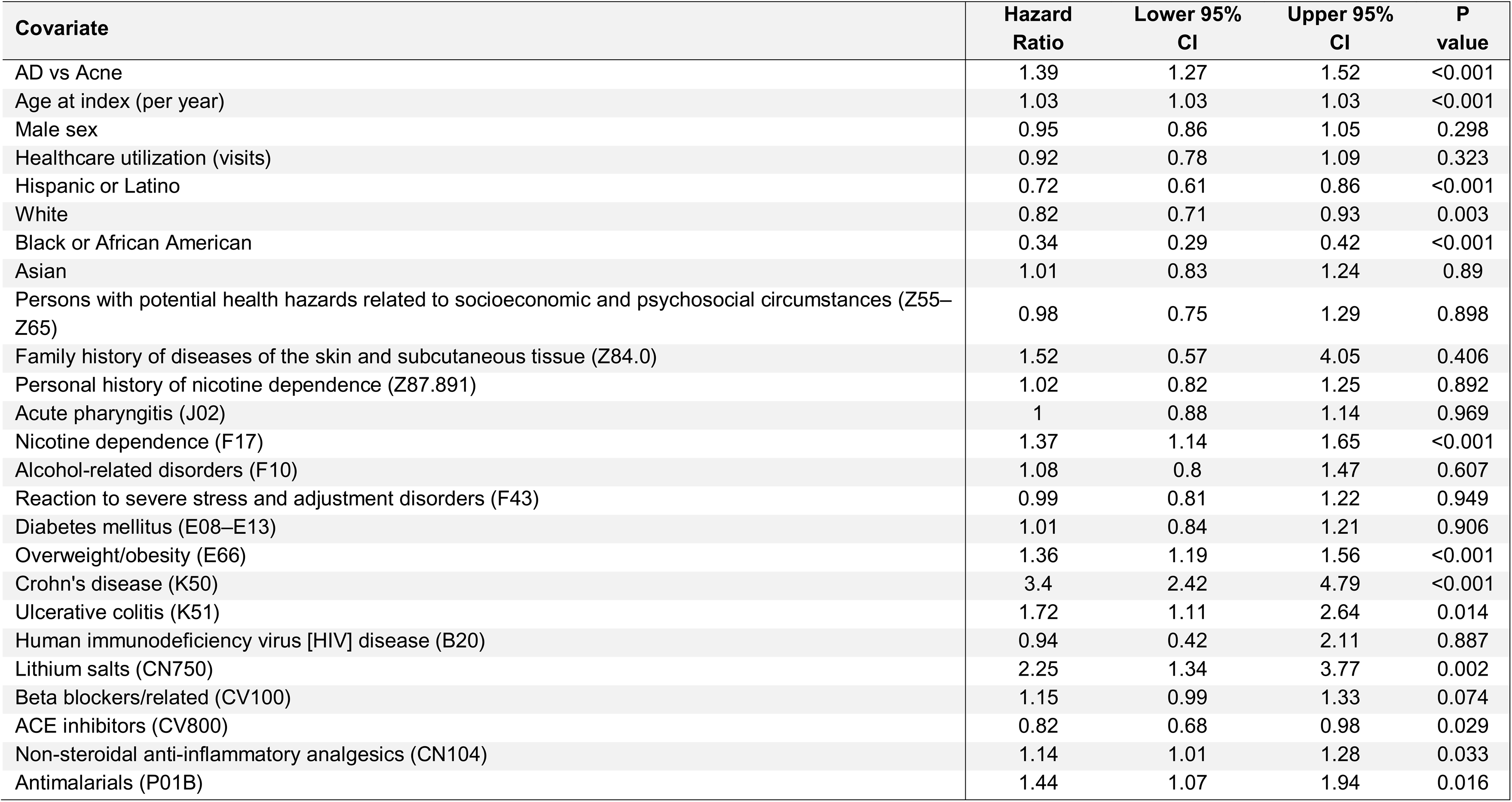
Multivariable Cox proportional hazards model for incident psoriasis vulgaris in patients with atopic dermatitis versus acne controls. Shown are results from a multivariable Cox proportional hazards regression model assessing incident psoriasis vulgaris in patients with atopic dermatitis compared with acne controls. The model included 351,074 patients with atopic dermatitis and 334,717 patients with acne and used a follow-up period of 2,595 days. Estimates are presented as hazard ratios with 95% confidence intervals. The model was adjusted for the covariates shown in the table.

Control outcome analyses supported the analytic framework. The positive control (topical betamethasone) was consistently elevated in patients with AD across the primary analysis, sensitivity analyses, and Cox model triangulation. Among negative controls, appendectomy remained null throughout Kaplan–Meier analyses. Presbycusis was null in the primary analysis, most sensitivity analyses, and Cox model triangulation, but showed an association in the acne comparator analysis. Burns and corrosions showed only small, inconsistent associations (Supplement Tables 3-4).

### Psoriasis risk in AD by systemic treatment exposure

In the primary analysis, 25,825 patients receiving biologic therapy and 5,457 receiving cvIS were identified before matching. Propensity score matching yielded 5,439 matched pairs. Before matching, cohorts differed modestly; after matching, all baseline covariates were well balanced (SMDs <0.10). Median follow-up was 596 days (biologic) and 601 days (cvIS), with interquartile ranges of 394 and 12 days, respectively. Full baseline characteristics are presented in Supplement Table 5.

Psoriasis was documented in 14 of 5,424 patients receiving biologic therapy (0.26%) versus 79 of 5,352 matched patients receiving cvIS (1.48%). Kaplan–Meier analysis demonstrated lower cumulative incidence in the biologic cohort (p<0.0001, HR, 0.20, 95%-CI, 0.11-0.35; Figure 2, Supplement Table 5).

**Figure 2.**
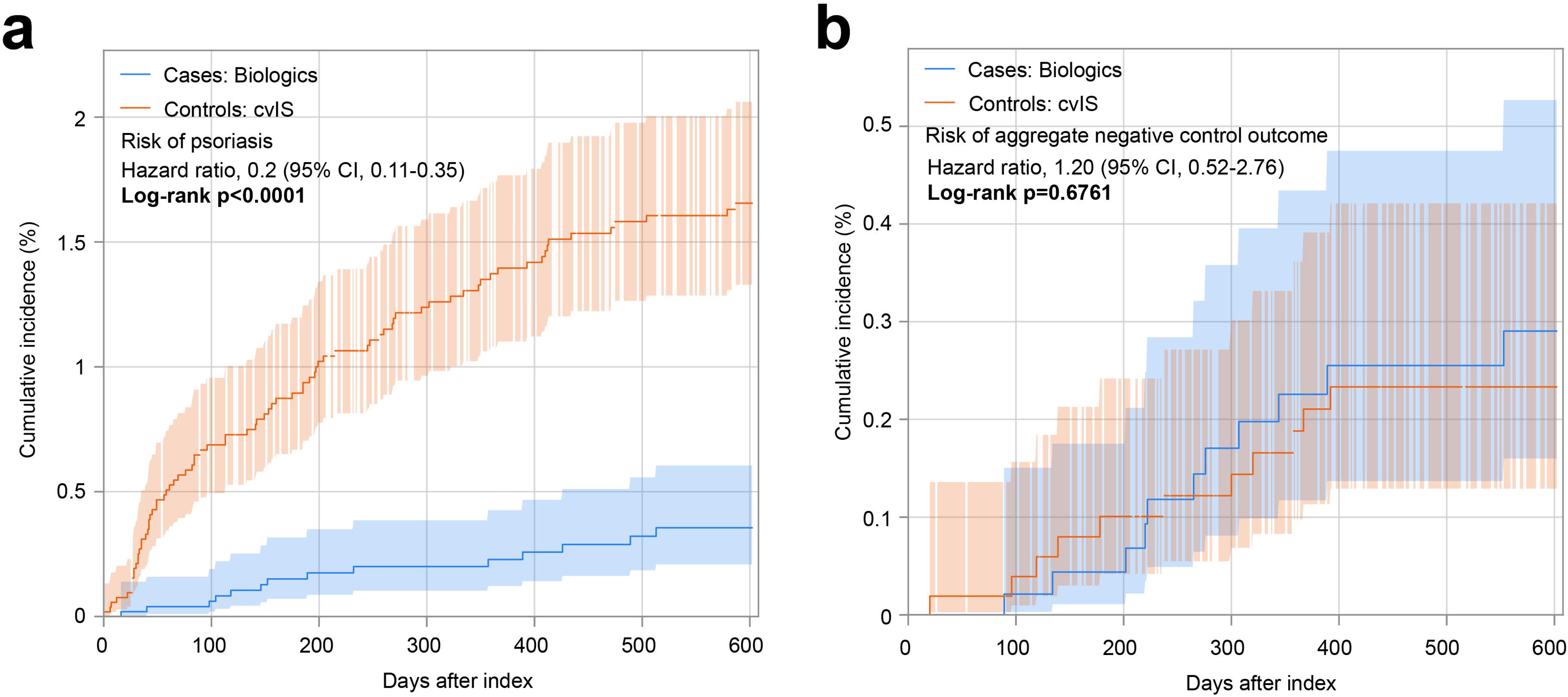
Kaplan-Meier analyses of psoriasis vulgaris and aggregate negative control outcomes in patients with atopic dermatitis (AD) treated with biologics versus conventional systemic immunosuppressants (cvIS). (**a**) Cumulative incidence of psoriasis vulgaris. Biologic therapy was associated with a lower cumulative incidence of psoriasis vulgaris than cvIS (HR, 0.20; 95% CI, 0.11–0.35; log-rank p<0.0001). (**b**) Cumulative incidence of the aggregate negative control outcome. No association was observed between treatment group and the aggregate negative control outcome (HR, 1.20; 95% CI, 0.52–2.76; log-rank p=0.6761). Shaded areas indicate 95% confidence intervals. Curves are displayed according to Kaplan-Meier convention, with the last observed survival probability carried forward until the next event or censoring.

The primary finding was directionally consistent across all evaluable sensitivity analyses, with hazard ratios consistently below 1.0 (range: 0.20–0.59, Supplement Table 5). The association persisted after restricting to patients with at least 6 months of baseline data (HR 0.32, 95%-CI, 0.18-0.57), using stricter AD definitions (HR 0.33,95%-CI, 0.19-0.59, and HR 0.36; 95%-CI, 0.23-0.56), among patients aged 18 years or older (HR 0.40, 95%-CI, 0.26-0.61), for dupilumab versus cvIS specifically (HR 0.35, 95%-CI, 0.22-0.54), and when the index event was temporally anchored on treatment initiation (HR 0.35, 95%-CI, 0.22-0.54). After applying a 3-month lag time, the association remained directionally consistent but was only nominally significant (HR 0.59, 95%-CI, 0.36-0.96; p=0.032). Analyses excluding pre-existing inflammatory skin diseases and those restricted to patients younger than 18 years were not evaluable due to insufficient outcome events. Multivariable Cox model triangulation confirmed the association (HR 0.28, 95%-CI, 0.22-0.37, p<0.001, Table 2).

**Table 2.** Multivariable Cox proportional hazards model for incident psoriasis vulgaris in patients treated with biologics versus conventional systemic immunosuppressants. Shown are results from a multivariable Cox proportional hazards regression model evaluating incident psoriasis vulgaris in patients treated with biologics compared with those treated with conventional systemic immunosuppressants (cvIS). The analysis included 27,407 patients receiving biologics and 5,940 receiving cvIS, with a follow-up of 601 days. Hazard ratios are presented with 95% confidence intervals. The model was adjusted for the covariates shown in the table.

| Covariate | Hazard Ratio | Lower 95% CI | Upper 95% CI | P value |
| --- | --- | --- | --- | --- |
| Biologic vs cvIS | 0.28 | 0.22 | 0.37 | <0.001 |
| Age at index (per year) | 1.02 | 1.01 | 1.02 | <0.001 |
| Male sex | 0.86 | 0.66 | 1.13 | 0.276 |
| Healthcare utilization (visits) | 1.26 | 0.52 | 3.09 | 0.611 |
| Hispanic or Latino | 0.94 | 0.55 | 1.6 | 0.813 |
| White | 1.11 | 0.74 | 1.66 | 0.616 |
| Black or African American | 0.36 | 0.2 | 0.66 | 0.001 |
| Asian | 1.76 | 1.09 | 2.84 | 0.021 |
| Persons with potential health hazards related to socioeconomic and psychosocial circumstances (Z55–Z65) | 0.88 | 0.45 | 1.71 | 0.695 |
| Family history of diseases of the skin and subcutaneous tissue (Z84.0) | 0.66 | 0.16 | 2.69 | 0.562 |
| Personal history of nicotine dependence (Z87.891) | 1.17 | 0.76 | 1.8 | 0.485 |
| Acute pharyngitis (J02) | 0.84 | 0.54 | 1.3 | 0.439 |
| Nicotine dependence (F17) | 1.63 | 1.07 | 2.51 | 0.025 |
| Alcohol-related disorders (F10) | 1.31 | 0.68 | 2.51 | 0.415 |
| Reaction to severe stress and adjustment disorders (F43) | 1.19 | 0.72 | 1.97 | 0.489 |
| Diabetes mellitus (E08–E13) | 1.08 | 0.69 | 1.69 | 0.737 |
| Overweight/obesity (E66) | 1.45 | 1 | 2.1 | 0.047 |
| Crohn's disease (K50) | 1.17 | 0.28 | 4.84 | 0.827 |
| Ulcerative colitis (K51) | 0.5 | 0.07 | 3.7 | 0.498 |
| Human immunodeficiency virus [HIV] disease (B20) | 0.86 | 0.12 | 6.24 | 0.881 |
| Lithium salts (CN750) | 1.03 | 0.14 | 7.54 | 0.974 |
| Beta blockers/related (CV100) | 0.96 | 0.65 | 1.42 | 0.835 |
| ACE inhibitors (CV800) | 0.91 | 0.58 | 1.43 | 0.677 |
| Non-steroidal anti-inflammatory analgesics (CN104) | 1.16 | 0.84 | 1.6 | 0.36 |
| Antimalarials (P01B) | 0.14 | 0.02 | 1.03 | 0.054 |

Negative control outcome analyses supported the analytic framework. In Kaplan–Meier analyses, the composite negative control outcome was evaluable only in the primary analysis and the sensitivity analysis with temporally anchored treatment exposure - both yielded null findings. Remaining sensitivity analyses were not evaluable due to insufficient outcome events (Supplement Table 5). Multivariable Cox model triangulation was consistent (HR 1.01, 95%-CI, 0.57-1.79; p=0.97; Supplement Table 6).

## Discussion

This large-scale, preregistered retrospective cohort study addressed two distinct questions. First, whether AD itself confers increased psoriasis risk - and second whether the choice of systemic treatment modifies that risk. Regarding the first question: AD was associated with a 1.4- to 4-fold increased risk of subsequent psoriasis compared with matched non-AD controls. Regarding the second: among patients with AD receiving systemic therapy, biologic treatment was associated with a 65–80% lower psoriasis risk compared with cvIS.

Turning to the second study question, whether treatment modifies psoriasis risk, the findings run counter to the prevailing concern that type 2-targeted biologics trigger psoriasis. Prior epidemiological studies have reported conflicting results, with some suggesting bidirectional associations and others supporting mutual exclusivity (7–9). The present study, including over 300,000 matched pairs with extended follow-up, provides robust evidence that AD confers increased psoriasis risk in routine clinical care.

The finding that biologic therapy was associated with reduced psoriasis risk compared with cvIS contrasts with recent reports suggesting that type 2-targeted biologics may unmask or trigger psoriasis (13–15). Furthermore, a recent large-scale retrospective cohort study reported a 58% increased psoriasis risk with dupilumab compared with other systemic agents (16). However, the study design may have been susceptible to bias, as it was not preregistered, did not tightly anchor treatment initiation to AD diagnosis, and was restricted to adults. An Italian study had also documented psoriasis in 1.3% of dupilumab-treated and 2.1% of tralokinumab-treated AD patients in a multicenter retrospective cohort explicitly designed to capture psoriasis outcomes (14) -potentially explaining the higher incidence compared with 0.26% observed here. Importantly, neither study included a cvIS comparator, precluding direct assessment of whether psoriasis risk differs by treatment class. The present study addresses this gap and suggests that when biologics are compared with cvIS in a matched framework, biologic therapy is associated with lower, not higher, psoriasis risk.

Notably, the absolute psoriasis incidence differed substantially across cohorts in a pattern potentially consistent with AD severity. In the overall AD versus non-AD comparison, which predominantly captures mild AD managed without systemic therapy, risk for a subsequent psoriasis diagnosis was 0.31%. In contrast, among patients with moderate-to-severe AD requiring systemic treatment, psoriasis incidence was 1.48% in those receiving cvIS - but only 0.26% in those receiving biologics. This gradient suggests that psoriasis risk may scale with AD severity – as observed for cutaneous T cell lymphoma risk in AD (20). If confirmed, this would imply that psoriasis emerging during AD treatment reflects disease-associated risk rather than a treatment-specific effect, with biologics potentially mitigating rather than exacerbating this risk.

Several design features support the validity of these findings. Control outcome analyses behaved largely as expected: the positive control (topical betamethasone) was consistently elevated in AD patients, while negative controls (appendectomy, presbycusis, burns/corrosions) were predominantly null. The study employed a stepwise approach to enhance transparency and minimize analytic flexibility: exploratory scoping analyses informed feasibility, followed by a formal design freeze, preregistration on the Open Science Framework, and confirmatory execution. This framework, herein termed “design freeze”, represents an adaptation of preregistration principles to federated EHR platforms where preliminary feasibility assessment is operationally necessary (17). Additionally, treatment cohorts were temporally anchored to mitigate era effects: eligibility began January 2016, approximately one year before dupilumab’s FDA approval for AD (March 2017). This design was necessary because prescribing patterns shifted substantially following biologic approval, which would otherwise limit temporal overlap between treatment groups and compromise comparability. Scoping analyses confirmed sufficient overlap in prescribing patterns during the study period (Figure 3).

**Figure 3.**
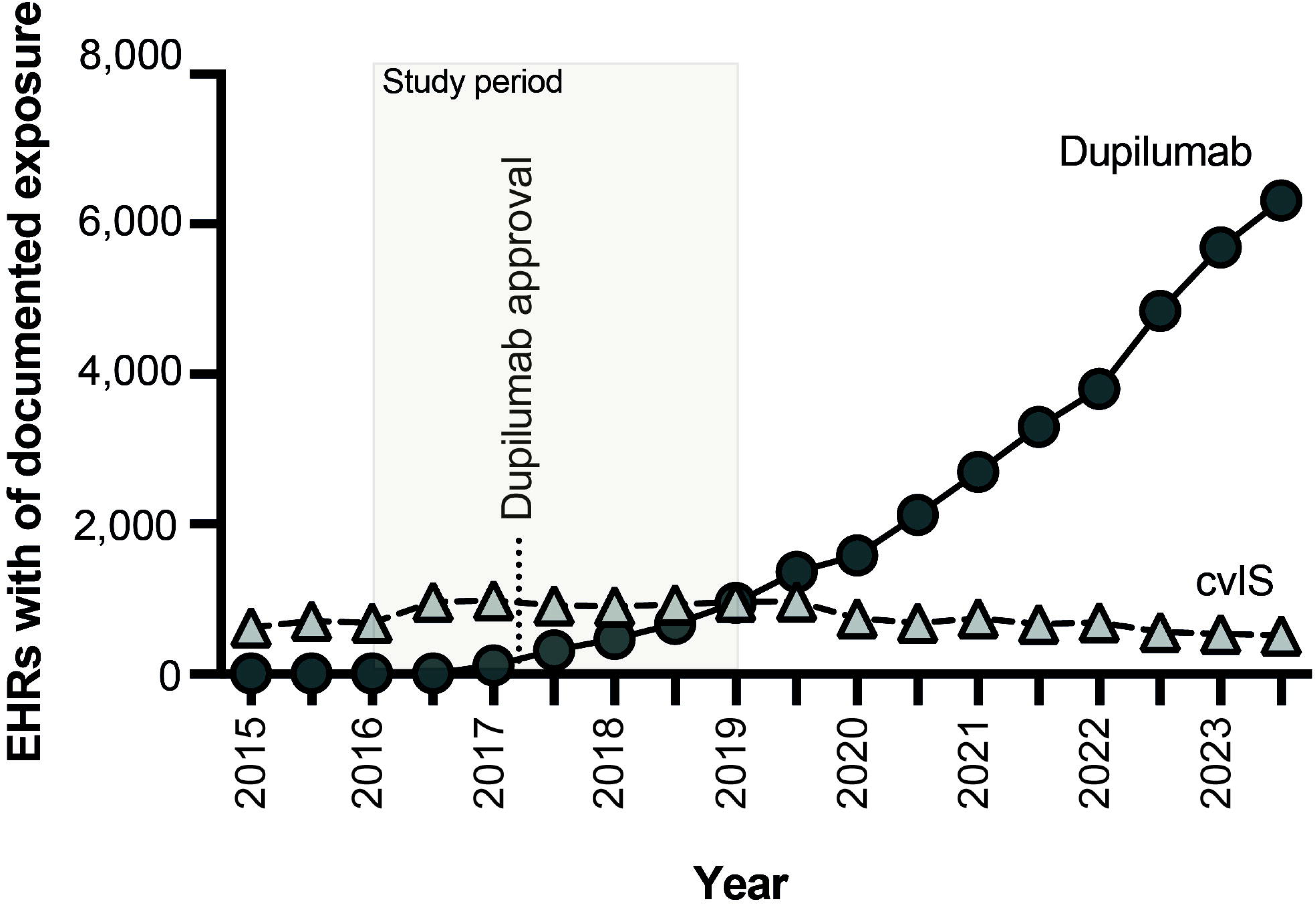
Temporal prescribing patterns of systemic therapies for atopic dermatitis in the TriNetX network. The number of patients with documented exposure to dupilumab and conventional systemic immunosuppressants (cvIS) was quantified in 6-month intervals across a 7-year window spanning 3 years before and 4 years after dupilumab approval for AD. Although the primary treatment analysis included tralokinumab and lebrikizumab alongside dupilumab in the biologic cohort, prescribing numbers for these agents were negligible during the study period; dupilumab therefore serves as the representative biologic agent in this figure. Based on the prespecified threshold for feasible comparative analyses, only dupilumab and cvIS showed sufficient temporal overlap and cohort size for confirmatory comparison. Cohort definitions otherwise mirrored the primary treatment analysis but did not exclude outcomes recorded before index. The shaded area indicates the study period used for the confirmatory treatment comparison, and the dotted vertical line marks the time when the first biologic (dupilumab) was approved for AD.

This study has several limitations inherent to retrospective EHR-based research. Residual confounding cannot be excluded despite propensity score matching for multiple covariates; unmeasured factors such as AD severity, disease duration, or prior treatment history may influence both treatment selection and psoriasis risk. Coding misclassification is possible, as diagnoses were ascertained from ICD-10-CM codes without clinical adjudication, and uncertainty regarding the exact treatment indication cannot be fully resolved, although requiring an AD diagnosis within one month of treatment initiation was intended to mitigate this concern. The observed psoriasis incidence of 0.08% among non-AD controls over 2,595 days corresponds to approximately 11.3 cases per 100,000 person-years, which is lower than expected population-based rates in children (approximately 33–41 per 100,000 person-years) (21). This discrepancy may in part reflect the constraints of EHR-based outcome ascertainment, including incomplete capture of diagnoses made outside the contributing healthcare network. The intention-to-treat–like framework, while reducing selection bias from treatment switching, may not fully capture treatment trajectories including discontinuation, dose changes, or sequential therapy. Proportionality assumptions were occasionally violated in Kaplan–Meier analyses. However, Cox model triangulation yielded directionally consistent findings, supporting the robustness of the primary conclusions. Additionally, although treatment cohorts were temporally anchored to overlap prescribing periods, residual era effects cannot be fully excluded; biologics and cvIS were still predominantly prescribed in different calendar periods, which may have introduced unmeasured confounding related to evolving clinical practice, patient selection, or concomitant treatment patterns. Finally, as an observational study, causal inference is not possible; the observed associations may reflect disease severity, healthcare utilization patterns, or other factors rather than direct biological effects of AD or its treatments on psoriasis development.

Taking the retrospective nature of this study into account, several clinical implications merit consideration. First, AD was associated with an increased risk of subsequent psoriasis. However, given the low absolute incidence of 0.31% over approximately seven years, these findings do not support routine screening for psoriasis in all patients with AD. Rather, they support clinical awareness that psoriasis may emerge in a subset of patients over time – particularly those with moderate to severe AD. Second, biologic treatment was not associated with increased psoriasis risk. Conversely, it was associated with a lower risk than cvIS. These data may inform treatment counselling and provide reassurance regarding the psoriasis risk profile of type 2-targeted biologics.

## Supporting information

STROBE

Detailed Maehtods

## Data Availability

All data produced in the present work are contained in the manuscript

## Acknowledgments

The authors thank Friederike Ubeing (University Hospital Schleswig-Holstein, Kiel, Germany) for her support in facilitating access to the TriNetX platform. During the preparation of this manuscript, the authors used ChatGPT (GPT-4o, OpenAI) for two purposes. First, to assist with the extraction and organization of data from the TriNetX platform; all extracted data were systematically verified for accuracy against the primary data by the authors prior to use. Second, to assist with language editing of selected manuscript passages to improve readability, using prompts of the nature of “please improve the scientific writing of the following text.” No AI assistance was used for statistical analysis, interpretation of results, or the drawing of scientific conclusions. All AI-assisted content was reviewed, edited, and approved by the authors, who take full responsibility for the integrity and accuracy of the published work.

This work was supported by the Cluster of Excellence *Precision Medicine in Chronic Inflammation* (EXC 2167), The Collaborative Research Center *Pathomechanisms of Antibody-mediated Autoimmunity* (SFB 1526), Individual Research Grant LU 877/25-1, all by the Deutsche Forschungsgemeinschaft, and the Schleswig-Holstein Excellence-Chair Program from the State of Schleswig-Holstein. PC was supported by Region Stockholm, Karolinska Institutet, Hudfonden, the Swedish Society for Dermatology and Venereology, and the Tore Nilson Foundation.

