## Supplementary material for "Incident psoriasis in atopic dermatitis: A large-scale cohort study of disease- and treatment-associated risks": STROBE

STROBE Statement—Checklist of items that should be included in reports of ***cohort studies***

|  | Item No | Recommendation |
| --- | --- | --- |
| **Title and abstract** | 1 | (*a*) Indicate the study’s design with a commonly used term in the title or the abstract Reported. The title identifies the study as “A large-scale cohort study,” and the abstract states that this was a preregistered retrospective cohort study using propensity score–matched analyses |
|  |  | (*b*) Provide in the abstract an informative and balanced summary of what was done and what was found Reported. The abstract summarizes background, objective, methods, results, limitations, and conclusion, including both aims, matching strategy, sensitivity analyses, model triangulation, and the main findings |
| Introduction | | |
| Background/rationale | 2 | Explain the scientific background and rationale for the investigation being reported Reported. The Introduction reviews the immunologic relationship between atopic dermatitis and psoriasis, conflicting epidemiologic and genetic evidence, and concern regarding biologics, especially dupilumab, as possible triggers of psoriasis. |
| Objectives | 3 | State specific objectives, including any prespecified hypotheses Reported. The Introduction states 2 aims: (1) whether AD is associated with increased incident psoriasis risk compared with matched non-AD controls, and (2) whether psoriasis risk differs according to systemic treatment exposure, particularly biologics versus conventional systemic immunosuppressants. The study is explicitly described as preregistered |
| Methods | | |
| Study design | 4 | Present key elements of study design early in the paper Reported. Early Methods describe a propensity score–matched retrospective cohort study using the TriNetX US Collaborative Network, with exploratory scoping, design freeze, preregistration, and confirmatory execution |
| Setting | 5 | Describe the setting, locations, and relevant dates, including periods of recruitment, exposure, follow-up, and data collection Reported. The setting is the TriNetX US Collaborative Network. Relevant dates are provided for index-event windows, treatment exposure windows, follow-up durations, design freeze, preregistration date, and confirmatory execution dates. |
| Participants | 6 | (*a*) Give the eligibility criteria, and the sources and methods of selection of participants. Describe methods of follow-up Reported. Eligibility criteria and cohort construction are described in the manuscript and in detail in Supplement Tables 1 and 2. Aim 1 defines AD cases and Z00 controls; Aim 2 defines biologic and cvIS treatment cohorts with AD required within 1 month before treatment. Follow-up began 1 day after index and was fixed at 2,595 days for Aim 1 and 601 days for Aim 2. |
|  |  | (*b*) For matched studies, give matching criteria and number of exposed and unexposed Reported. Matching used 1:1 greedy nearest-neighbor propensity score matching with caliper 0.1 SD. Numbers before and after matching are given for both aims: Aim 1, 351,070 AD and 6,888,769 controls before matching, 351,069 matched pairs after matching; Aim 2, 25,825 biologic and 5,457 cvIS before matching, 5,439 matched pairs after matching. |
| Variables | 7 | Clearly define all outcomes, exposures, predictors, potential confounders, and effect modifiers. Give diagnostic criteria, if applicable Reported. Primary outcome, control outcomes, treatment exposures, covariates, and age-stratified analyses are defined in the manuscript and detailed Methods, with coding logic in Supplement Tables 1 and 2. ICD-10-CM, CPT, SNOMED, and RxNorm code-based definitions are specified. |
| Data sources/ measurement | 8* | For each variable of interest, give sources of data and details of methods of assessment (measurement). Describe comparability of assessment methods if there is more than one group Reported. All variables were derived from TriNetX federated EHR data using coded diagnoses, procedures, and medication records. The same platform and coding framework were used across exposed and comparator groups; supplementary cohort-definition tables provide detailed coding and temporal rules |
| Bias | 9 | Describe any efforts to address potential sources of bias Reported. The manuscript describes several measures to address bias: scoping without confirmatory inference, design freeze, preregistration, active comparator design in Aim 2, tight anchoring of AD diagnosis to treatment, exclusion of prevalent outcomes, propensity score matching, prespecified sensitivity analyses, model triangulation, and positive/negative control outcomes. Limitations such as residual confounding, coding misclassification, treatment-indication uncertainty, and observational design are also discussed. |
| Study size | 10 | Explain how the study size was arrived at Reported. Study size was determined by the number of eligible patients available in the TriNetX network under the prespecified cohort definitions. For Aim 2, scoping analyses used a predefined threshold of at least 1,000 patients for feasible treatment comparisons, leading to retention of biologic/cvIS and exclusion of smaller JAK inhibitor and IL-13 inhibitor cohorts. |
| Quantitative variables | 11 | Explain how quantitative variables were handled in the analyses. If applicable, describe which groupings were chosen and why Reported in part. Continuous variables such as age and healthcare utilization were modeled quantitatively; age-stratified sensitivity analyses used predefined age groups (<18 years and ≥18 years). Follow-up was fixed based on scoping. Additional explanation of why these groupings were chosen is given for sensitivity analyses and feasibility |
| Statistical methods | 12 | (*a*) Describe all statistical methods, including those used to control for confounding Reported. Statistical methods included propensity score matching by logistic regression, greedy nearest-neighbor matching, standardized mean differences for balance, Kaplan–Meier analyses with log-rank tests, hazard ratios with 95% CIs, multivariable Cox proportional hazards model triangulation, and Bonferroni correction. |
|  |  | (*b*) Describe any methods used to examine subgroups and interactions Reported. Prespecified subgroup/sensitivity analyses included lag-time analyses, minimum baseline window, stricter AD definitions, exclusion analyses, age stratification, alternative comparator, dupilumab-only comparison, and temporally anchored treatment indexing. No formal interaction terms were reported. |
|  |  | (*c*) Explain how missing data were addressed Reported. Missing covariate values were retained in all analyses; in propensity score matching, missingness was treated as a separate category, and in Cox models missing values were likewise retained. |
|  |  | (*d*) If applicable, explain how loss to follow-up was addressed Reported in part. Follow-up was administratively fixed at the shorter common maximum follow-up identified in scoping (2,595 days for Aim 1; 601 days for Aim 2), thereby standardizing observation windows across cohorts. The study used EHR follow-up within the TriNetX network rather than direct participant follow-up. |
|  |  | (*e*) Describe any sensitivity analyses Reported. Eight prespecified sensitivity analyses were performed for Aim 1 and nine for Aim 2, as detailed in the manuscript and detailed Methods, including lag-time, baseline-window, stricter AD definitions, age stratification, comparator changes, and treatment-anchored definitions. |
| Results | | |
| Participants | 13* | (a) Report numbers of individuals at each stage of study—eg numbers potentially eligible, examined for eligibility, confirmed eligible, included in the study, completing follow-up, and analysed Reported in part. Numbers before and after matching are presented for both aims, and relevant treatment-cohort feasibility counts from scoping are given. The manuscript reports final analyzed cohort sizes and evaluability of sensitivity analyses. |
|  |  | (b) Give reasons for non-participation at each stage Not directly applicable in the traditional prospective sense. This was a retrospective EHR study using coded inclusion and exclusion criteria rather than participant recruitment. Reasons for exclusion are described through cohort-definition rules and prior-outcome/treatment exclusions. |
|  |  | (c) Consider use of a flow diagram Not done |
| Descriptive data | 14* | (a) Give characteristics of study participants (eg demographic, clinical, social) and information on exposures and potential confounders Reported. Baseline characteristics before and after matching are summarized in the Results and in Supplement Tables 3 and 5, including age, sex, race/ethnicity/ancestry categories, clinical covariates, and medication exposures. |
|  |  | (b) Indicate number of participants with missing data for each variable of interest Partly reported. The manuscript states how missing data were handled, but does not provide a dedicated variable-by-variable table of missingness in the main text. Missingness is retained within TriNetX analyses and matching/Cox procedures. |
|  |  | (c) Summarise follow-up time (eg, average and total amount) Reported. Median follow-up and interquartile ranges are given for both aims; fixed follow-up windows are also stated |
| Outcome data | 15* | Report numbers of outcome events or summary measures over time Reported. The manuscript gives event counts, risks, hazard ratios, and Kaplan–Meier results for psoriasis and control outcomes in both aims; additional results are summarized in supplementary tables. |
| Main results | 16 | (*a*) Give unadjusted estimates and, if applicable, confounder-adjusted estimates and their precision (eg, 95% confidence interval). Make clear which confounders were adjusted for and why they were included Reported. Main matched analyses provide hazard ratios with 95% CIs, and multivariable Cox triangulation provides adjusted estimates with listed covariates in Tables 1 and 2. The rationale for covariate selection is described in detailed Methods, including clinical relevance and scoping feasibility. |
|  |  | (*b*) Report category boundaries when continuous variables were categorized Reported where applicable. Age stratification is reported as <18 years and ≥18 years. Baseline-window and lag-time sensitivity analyses are also explicitly defined |
|  |  | (*c*) If relevant, consider translating estimates of relative risk into absolute risk for a meaningful time period Reported in part. Absolute risks are provided for primary outcomes and controls. The Discussion additionally translates the non-AD control psoriasis incidence to approximately 11.3 cases per 100,000 person-years. |
| Other analyses | 17 | Report other analyses done—eg analyses of subgroups and interactions, and sensitivity analyses Reported. The manuscript reports prespecified sensitivity analyses, age-stratified analyses, dupilumab-specific comparison, temporally anchored treatment analysis, control outcome analyses, and multivariable Cox triangulation. |
| Discussion | | |
| Key results | 18 | Summarise key results with reference to study objectives Reported. The Discussion opens by summarizing the 2 principal findings directly aligned with the 2 study aims. |
| Limitations | 19 | Discuss limitations of the study, taking into account sources of potential bias or imprecision. Discuss both direction and magnitude of any potential bias Reported. The Discussion addresses residual confounding, coding misclassification, treatment-indication uncertainty, low control incidence, intention-to-treat–like exposure modeling, proportionality concerns, sparse events in some analyses, and inability to infer causality. Direction and possible magnitude are discussed qualitatively. |
| Interpretation | 20 | Give a cautious overall interpretation of results considering objectives, limitations, multiplicity of analyses, results from similar studies, and other relevant evidence Reported. The Discussion interprets the findings in light of prior epidemiologic, genetic, and pharmacovigilance literature, contrasts them with recent TriNetX and Italian studies, and remains cautious about causality and treatment-associated interpretation. |
| Generalisability | 21 | Discuss the generalisability (external validity) of the study results Reported. The manuscript notes use of a large US TriNetX network and discusses generalizability, including broader relevance beyond adults due to inclusion of pediatric patients, while acknowledging limitations of EHR-based ascertainment and network coverage. |
| Other information | | |
| Funding | 22 | Give the source of funding and the role of the funders for the present study and, if applicable, for the original study on which the present article is based Reported. Funding sources are explicitly listed in the manuscript. The paper also includes disclosure statements. No direct funder role in study conduct is described in the current text |

*Give information separately for exposed and unexposed groups.

**Note:** An Explanation and Elaboration article discusses each checklist item and gives methodological background and published examples of transparent reporting. The STROBE checklist is best used in conjunction with this article (freely available on the Web sites of PLoS Medicine at http://www.plosmedicine.org/, Annals of Internal Medicine at http://www.annals.org/, and Epidemiology at http://www.epidem.com/). Information on the STROBE Initiative is available at http://www.strobe-statement.org.
