## Supplementary material for "Incident psoriasis in atopic dermatitis: A large-scale cohort study of disease- and treatment-associated risks": Detailed Maehtods

**Detailed materials and methods**

**Study design and data source**

A propensity score-matched (PSM) retrospective cohort study was conducted using the US Collaborative Network of the federated electronic health record (EHR) platform TriNetX (following established procedures (1). The study comprised an exploratory scoping phase to assess feasibility (including cohort definitions, the number of covariates accommodated by the propensity-score matching (PSM), and sufficiency of outcome events), followed by a design freeze, preregistration (Open Science Framework: (<https://osf.io/8f2s4/registrations>), registered March 26, 2026), and confirmatory execution (performed March 27-28, 2026).

Analyses were performed within the TriNetX platform using federated queries, with only aggregated results returned to investigators. The network was selected for its large EHR volume, detailed covariate documentation, and representation of the US healthcare-seeking population (2,3). At the time of analysis, 65-66 of a total of 66 Healthcare Organizations within the network provided data from over 110 million electronic healthcare records (EHRs). The study evaluated whether AD was associated with an increased risk of incident psoriasis and whether psoriasis risk differed according to treatment exposure among patients with AD. Reporting followed the STROBE guidelines (4). The study evaluated whether AD was associated with an increased risk of incident psoriasis and whether psoriasis risk differed according to treatment exposure among patients with AD. The analysis used de-identified secondary data and involved no interaction with human participants. Data were de-identified according to §164.514(a) of the HIPAA Privacy Rule. The TriNetX platform operates under a federated governance framework compliant with HIPAA, the General Data Protection Regulation (GDPR), and the Lei Geral de Proteção de Dados (LGPD); therefore, institutional review board approval was not required.

**Preliminary scoping analysis and design finalization**

Before formal execution of the pre-registered analyses, a scoping analysis was conducted for both study questions using the logic of the planned primary analyses. In this phase, outcomes were counted beginning 1 day after the index event without additional follow-up restrictions to verify technical feasibility of cohort definitions, covariate retrieval, comparator construction, and outcome ascertainment within TriNetX. This step served only to confirm feasibility and operational validity of the intended design and was not used for confirmatory inference. For Aim 2, the scoping analysis revealed that only the biologic versus cvIS and dupilumab versus cvIS comparisons yielded sufficient sample sizes for comparative analysis; Janus kinase inhibitor (n=983) and IL-13 inhibitor (n=430) cohorts fell below the prespecified threshold of 1,000 patients after cohort construction and were therefore not taken forward. After review of the scoping outputs, the final analytic framework was fixed at a design freeze point (March 26th, 2026), defined as the transition from exploratory setup to locked confirmatory design. At this point, cohort definitions, covariates, outcomes, comparators, and sensitivity analyses were finalized and no longer modified on the basis of observed data patterns. Once preregistered, the full study was executed.

**Study population and index events**

Aim 1. Cases were defined as patients with a first recorded diagnosis of atopic dermatitis (ICD-10-CM L20) between 1 January 2016 and 31 December 2018. Controls were defined as patients with a first recorded general health examination encounter (Z00) during the same period. For both cohorts, the index date was the date of the first eligible record of the respective index event. At data retrieval, patients with recorded psoriasis, burns and corrosions of external body surface, appendectomy, presbycusis, topical betamethasone exposure, or prior atopic dermatitis on or before the index date were excluded. Detailed cohort definitions and index event specifications are provided in Supplementary Table 1.

Aim 2. Treatment cohorts were defined from first eligible systemic treatment exposure on or after 1 January 2017. This design was chosen in part to minimize distortion from treatment-era effects, which may influence comparative estimates when biologics and cvIS are assessed across substantially different prescribing periods. The primary exposed cohort consisted of patients receiving dupilumab, tralokinumab, or lebrikizumab, and the comparator cohort of patients receiving methotrexate, azathioprine, mycophenolate mofetil, or ciclosporin. In both groups, the index date was the date of first eligible treatment exposure. Exposure was defined at first eligible treatment record, with follow-up proceeding irrespective of subsequent treatment changes, consistent with an intention-to-treat–like framework. To align treatment exposure with indication, a recorded diagnosis of atopic dermatitis (L20) was required within 1 month on or before treatment initiation. To ensure treatment-naïve comparisons, patients in the biologic cohort with prior exposure to methotrexate, azathioprine, mycophenolate mofetil, ciclosporin, or Janus kinase inhibitors (abrocitinib, upadacitinib, baricitinib) on or before index were excluded. Likewise, patients in the cvIS cohort with prior exposure to dupilumab, tralokinumab, lebrikizumab, or Janus kinase inhibitors on or before index were excluded. Outcomes recorded on or before the index event were excluded at data retrieval. Detailed cohort definitions and index event specifications are provided in Supplementary Table 2.

To contextualize the treatment comparison against temporal prescribing patterns, scoping analyses quantified the number of patients receiving each eligible therapy in 6-month intervals across a 6-year window spanning 3 years before and 4 years after the year in which the respective drug was licensed for atopic dermatitis. Based on the prespecified threshold, comparative analyses were restricted to dupilumab and cvIS. Cohort definitions otherwise mirrored the primary analysis, but without exclusion of outcomes recorded before index.

**Outcomes**

Outcomes were defined by ICD-10-CM, SNOMED or CPT codes and included psoriasis vulgaris (L40.0), Burns and corrosions of external body surface (T20-T25), presbycusis (H91.1), and appendectomy (CPT 1014622, 44950, 44970, SNOMED 80146002) served as negative control outcomes across all analyses. Topical betamethasone (RxNorm 1514; topical product) served as a positive control outcome in Aim 1. Outcomes recorded before the start of follow-up were excluded at data retrieval. If the number of events was too low for individual negative control outcomes, a composite of all was used. Additional outcomes listed in the preregistration (atopic dermatitis, acne vulgaris, rosacea, hidradenitis suppurativa, vitiligo, alopecia areata, lichen planus, and depression) were not analyzed, as these entries were inadvertently carried over into the preregistration and were not part of the prespecified study objective addressed here. This did not affect cohort construction, outcome ascertainment for the reported endpoint, or the analyses presented in this manuscript.

**Covariates and propensity score matching**

Aim 1. Covariates for propensity score matching (PSM) were selected a priori based on clinical relevance and scoping feasibility. Because the scoping analysis showed that no more than 14 covariates could be accommodated in the TriNetX matching framework, the final propensity score model included age at index, female sex, White ancestry, Black or African American ancestry, socioeconomic and psychosocial circumstances (Z55-Z65), personal history of nicotine dependence (Z87.891), acute pharyngitis (J02), nicotine dependence (F17), alcohol-related disorders (F10), diabetes mellitus (E08-E13), overweight and obesity (E66), beta blockers/related (VA CV100), ACE inhibitors (VA CV800), and non-steroidal anti-inflammatory analgesics (VA CN104), all assessed up to 1 day before index. Propensity scores were estimated by logistic regression and cohorts matched 1:1 using greedy nearest-neighbor matching with a caliper of 0.1 pooled standard deviations of the logit of the propensity score; balance was assessed using standardized mean differences. Additional retrieved but unmatched covariates included male sex, Hispanic or Latino ancestry, Asian ancestry, family history of skin and subcutaneous tissue disease (Z84.0), reaction to severe stress and adjustment disorders (F43), Crohn's disease (K50), ulcerative colitis (K51), HIV disease (B20-B24), lithium salts (VA CN750), and antimalarials (VA AP101). Based on the scoping analysis, follow-up was fixed at 2,595 days.

Aim 2. Propensity score matching for Aim 2 followed the same approach as Aim 1, with all covariates included. Follow-up was fixed at 601 days based on the scoping analysis.

**Sensitivity analyses**

Aim 1. The primary analysis compared incident AD with non-AD controls. Follow-up began 1 day after the index event and was restricted to the maximum common fixed follow-up period supported by both cohorts, as determined in the scoping analysis (2,595 days). Thus, the prespecified follow-up window corresponded to the shorter of the two cohort-specific available follow-up durations identified during scoping and was applied uniformly to both cohorts and across all analyses. Prespecified sensitivity analyses included a 3-month lag-time analysis (S1), restriction to patients with at least 6 months of baseline observation (S2), and a stricter AD definition requiring two recorded diagnoses of atopic dermatitis (L20), with the first instance of AD occurring within 6 weeks to 12 months before any instance of AD (Group 1A); the index date remained the Group 1A event, so this definition did not introduce immortal time bias (S3). Alternatively, in S4, AD was defined using an alternative two-diagnosis logic requiring a second qualifying L20 diagnosis within 1 month on or after the first AD diagnosis; the corresponding logic was applied to controls. This design introduces immortal time bias because cohort qualification depends on a post-index event but was included for comparison with commonly used observational database definitions. Due to computational constraints related to cohort size, propensity score matching in S4 was limited to 13 covariates (excluding non-steroidal anti-inflammatory analgesics). In S5, diagnoses of dermatitis and eczema (L20-L30), papulosquamous disorders (L40-L45), and other disorders of the skin and subcutaneous tissue (L49-L50) recorded before index were excluded in both cases and controls, except for L20 in the AD group. In S6, incident acne vulgaris served as an active comparator cohort. Additional analyses were stratified by age, separately including patients aged under 18 years (S7) and 18 years or older (S8). Multivariable Cox proportional hazards model triangulation (S9) was prespecified but could not be executed due to computational constraints; the model failed to converge even when covariates were reduced to six. This was acknowledged in the preregistration as potentially problematic. As a post-hoc alternative, the non-AD control definition was modified by replacing general health examination encounters (Z00) with acne vulgaris (L70.0) as the index event.

Aim 2. The primary analysis compared treatment cohorts using follow-up beginning 1 day after the index event and uniformly capped at the maximum common follow-up duration available in both cohorts (601 days), corresponding to the shorter observable follow-up of either cohort identified during scoping. Prespecified sensitivity analyses included a 3-month lag-time analysis (S1), restriction to patients with at least 6 months of baseline observation before index (S2), and a stricter AD definition requiring two recorded diagnoses of atopic dermatitis (L20), with cohort entry anchored to the qualifying index event so that no immortal time bias was introduced (S3). In S4, the same alternative two-diagnosis AD definition as in Aim 1 was applied in both treatment groups, acknowledging the same immortal time bias considerations described above. In S5, the same exclusion of pre-index diagnoses as in Aim 1 was applied in both treatment groups. Additional analyses were stratified by age, separately including patients aged under 18 years (S6) and 18 years or older (S7). In S8, dupilumab alone was compared with conventional systemic immunosuppressants. JAK inhibitor comparison was prespecified but not performed as the cohort did not meet the minimum threshold of 1,000 patients. In S9, exclusion of outcomes prior to the index event was temporally anchored on applied treatments, as opposed to AD diagnosis. In S10, model triangulation was performed using multivariable Cox proportional hazards models (see below).

**Model triangulation**

Model triangulation was performed using multivariable Cox proportional hazards models in addition to propensity score–matched analyses. For both aims, the covariate set mirrored that used in Aim 2 propensity score matching, with female sex replaced by male sex as the reference category (TriNetX platform default). Aim 1: AD status was entered as the main predictor. Follow-up was fixed at 2,595 days, consistent with the primary Kaplan-Meier analysis. Outcomes included psoriasis vulgaris, topical betamethasone (positive control), and one negative control outcome (presbycusis). As anticipated in the preregistration, computational constraints prevented Cox model execution using the original non-AD control definition (Z00). As a post-hoc alternative, the control cohort was redefined using acne vulgaris (L70.0) as the index event. Aim 2: Treatment cohort (biologics versus cvIS) was entered as the main predictor. Follow-up was fixed at 601 days, consistent with the primary Kaplan-Meier analysis. Outcomes included psoriasis vulgaris and a composite negative control outcome (burns and corrosions, presbycusis, and appendectomy), as individual negative control outcomes had insufficient event counts for separate analysis.

**Missing data handling**

Missing covariate values were retained in all analyses. In propensity score matching, missingness was treated as a separate category. In multivariable Cox proportional hazards models, missing values were likewise retained.

**Statistical analysis**

Baseline characteristics were summarized before and after propensity score matching. Continuous variables were compared using t-tests and binary or categorical variables using z-tests; covariate balance was assessed using standardized mean differences. Time-to-event analyses were performed using the Kaplan-Meier method and compared using the log-rank test, with hazard ratios and 95% confidence intervals reported For Kaplan-Meier analyses, TriNetX reports a proportionality test including a chi-square statistic and p-value. Consistent with platform guidance, these were interpreted qualitatively rather than as strict inferential thresholds: larger chi-square values were taken to indicate less proportionality and smaller values greater proportionality. Bonferroni correction was applied for multiple testing across prespecified outcomes (Aim 1: α(adjust)=0.01; aim 2: α(adjust)=0.025).
