## Supplementary material for "Incident psoriasis in atopic dermatitis: A large-scale cohort study of disease- and treatment-associated risks": Sup Tab 1

**Supplement Table 1.** Cohort construction, index event definitions, and temporal rules for the primary and sensitivity analyses for Aim 1.

| **Primary analysis, S1** | | | | |
| --- | --- | --- | --- | --- |
| **Cohort** | **Query group** | **Definition** | **Codes** | **Temporal rule** |
| **Atopic dermatitis (cases)** | Group 1A | Index event: first recorded diagnosis of atopic dermatitis | ICD-10-CM: L20 – Atopic dermatitis | Event between 1 Jan 2016 and 31 Dec 2018 |
|  | Group 1B | Exclusion criteria prior to cohort entry | ICD-10-CM: L40 – Psoriasis; ICD-10-CM: T20–T25 – Burns and corrosions of external body surface, specified by site; CPT: 1014622, 44950, 44960, 44970 – Appendectomy; SNOMED: 80146002 – Appendectomy; ICD-10-CM: H91.1 – Presbycusis; RxNorm: 1514 – Betamethasone (topical product) | Any instance of Group 1B occurred on or before the first instance of Group 1A |
| **Non-atopic dermatitis controls** | Group 1A | Index event: encounter for general examination without complaint, suspected or reported diagnosis | ICD-10-CM: Z00 – Encounter for general examination without complaint, suspected or reported diagnosis | Event between 1 Jan 2016 and 31 Dec 2018 |
|  | Group 1B | Exclusion criteria prior to cohort entry | ICD-10-CM: L40 – Psoriasis; ICD-10-CM: T20–T25 – Burns and corrosions of external body surface, specified by site; CPT: 1014622, 44950, 44960, 44970 – Appendectomy; SNOMED: 80146002 – Appendectomy; ICD-10-CM: H91.1 – Presbycusis; RxNorm: 1514 – Betamethasone (topical product) | Any instance of Group 1B occurred on or before the first instance of Group 1A |

| **S2** | | | | |
| --- | --- | --- | --- | --- |
| **Cohort** | **Query group** | **Definition** | **Codes** | **Temporal rule** |
| **Atopic dermatitis (cases)** | Group 1A | Index event: first recorded diagnosis of atopic dermatitis | ICD-10-CM: L20 – Atopic dermatitis | Event between 1 Jan 2016 and 31 Dec 2018 |
|  | Group 1B | Exclusion criteria prior to cohort entry | ICD-10-CM: L40 – Psoriasis; ICD-10-CM: T20–T25 – Burns and corrosions of external body surface, specified by site; CPT: 1014622, 44950, 44960, 44970 – Appendectomy; SNOMED: 80146002 – Appendectomy; ICD-10-CM: H91.1 – Presbycusis; RxNorm: 1514 – Betamethasone (topical product) | Any instance of Group 1B occurred on or before the first instance of Group 1A |
|  | Group 2A | Atopic dermatitis diagnosis for patients meeting the minimum baseline requirement | ICD-10-CM: L20 – Atopic dermatitis | Any instance of Group 2B occurred at least 6 months before the first instance of Group 2A |
|  | Group 2B | Healthcare encounter establishing baseline availability | Visit | Must have occurred at least 6 months before the first instance of Group 2A |
| **Non-atopic dermatitis controls** | Group 1A | Index event: encounter for general examination without complaint, suspected or reported diagnosis | ICD-10-CM: Z00 – Encounter for general examination without complaint, suspected or reported diagnosis | Event between 1 Jan 2016 and 31 Dec 2018 |
|  | Group 1B | Exclusion criteria prior to cohort entry | ICD-10-CM: L40 – Psoriasis; ICD-10-CM: T20–T25 – Burns and corrosions of external body surface, specified by site; CPT: 1014622, 44950, 44960, 44970 – Appendectomy; SNOMED: 80146002 – Appendectomy; ICD-10-CM: H91.1 – Presbycusis; RxNorm: 1514 – Betamethasone (topical product) | Any instance of Group 1B occurred on or before the first instance of Group 1A |
|  | Group 2A | Control index event for patients meeting the minimum baseline requirement | ICD-10-CM: Z00 – Encounter for general examination without complaint, suspected or reported diagnosis | Any instance of Group 2B occurred at least 6 months before the first instance of Group 2A |
|  | Group 2B | Healthcare encounter establishing baseline availability | Visit | Must have occurred at least 6 months before the first instance of Group 2A |

| **S3** | | | | |
| --- | --- | --- | --- | --- |
| **Cohort** | **Query group** | **Definition** | **Codes** | **Temporal rule** |
| **Atopic dermatitis (cases)** | Group 1A | Index event: first recorded diagnosis of atopic dermatitis | ICD-10-CM: L20 – Atopic dermatitis | Event between 1 Jan 2016 and 31 Dec 2018 |
|  | Group 1B | Exclusion criteria prior to cohort entry | ICD-10-CM: L40 – Psoriasis; ICD-10-CM: T20–T25 – Burns and corrosions of external body surface, specified by site; CPT: 1014622, 44950, 44960, 44970 – Appendectomy; SNOMED: 80146002 – Appendectomy; ICD-10-CM: H91.1 – Presbycusis; RxNorm: 1514 – Betamethasone (topical product) | Any instance of Group 1B occurred on or before the first instance of Group 1A |
|  | Group 2A | Atopic dermatitis diagnosis meeting the stricter case-definition framework | CD-10-CM: L20 – Atopic dermatitis | The first instance of Group 2B occurred within 6 weeks and 1 year before any instance of Group 2A |
|  | Group 2B | Prior confirming diagnosis of atopic dermatitis | ICD-10-CM: L20 – Atopic dermatitis | Must have occurred within 6 weeks to 1 year before the qualifying Group 2A diagnosis |
| **Non-atopic dermatitis controls** | Group 1A | Index event: encounter for general examination without complaint, suspected or reported diagnosis | ICD-10-CM: Z00 – Encounter for general examination without complaint, suspected or reported diagnosis | Event between 1 Jan 2016 and 31 Dec 2018 |
|  | Group 1B | Exclusion criteria prior to cohort entry | ICD-10-CM: L40 – Psoriasis; ICD-10-CM: T20–T25 – Burns and corrosions of external body surface, specified by site; CPT: 1014622, 44950, 44960, 44970 – Appendectomy; SNOMED: 80146002 – Appendectomy; ICD-10-CM: H91.1 – Presbycusis; RxNorm: 1514 – Betamethasone (topical product) | Any instance of Group 1B occurred on or before the first instance of Group 1A |
|  | Group 2A | Control index event meeting the stricter temporal definition | ICD-10-CM: Z00 – Encounter for general examination without complaint, suspected or reported diagnosis | Any instance of Group 2B occurred within 6 weeks and 1 year before the first instance of Group 2A |
|  | Group 2B | Prior healthcare encounter | Visit | Must have occurred within 6 weeks to 1 year before the first instance of Group 2A |

| **S4** | | | | |
| --- | --- | --- | --- | --- |
| **Cohort** | **Query group** | **Definition** | **Codes** | **Temporal rule** |
| **Atopic dermatitis (cases)** | Group 1A | Index event: first recorded diagnosis of atopic dermatitis | ICD-10-CM: L20 – Atopic dermatitis | Event between 1 Jan 2016 and 31 Dec 2018 |
|  | Group 1B | Exclusion criteria prior to cohort entry | ICD-10-CM: L40 – Psoriasis; ICD-10-CM: T20–T25 – Burns and corrosions of external body surface, specified by site; CPT: 1014622, 44950, 44960, 44970 – Appendectomy; SNOMED: 80146002 – Appendectomy; ICD-10-CM: H91.1 – Presbycusis; RxNorm: 1514 – Betamethasone (topical product) | Any instance of Group 1B occurred on or before the first instance of Group 1A |
|  | Group 2A | Atopic dermatitis diagnosis meeting the two-code sensitivity definition | ICD-10-CM: L20 – Atopic dermatitis | Any instance of Group 2B occurred within 1 month on or after the first instance of Group 2A |
|  | Group 2B | Confirming second diagnosis of atopic dermatitis | ICD-10-CM: L20 – Atopic dermatitis | Must have occurred within 1 month on or after the first instance of Group 2A |
| **Non-atopic dermatitis controls** | Group 1A | Index event: encounter for general examination without complaint, suspected or reported diagnosis | ICD-10-CM: Z00 – Encounter for general examination without complaint, suspected or reported diagnosis | Event between 1 Jan 2016 and 31 Dec 2018 |
|  | Group 1B | Exclusion criteria prior to cohort entry | ICD-10-CM: L40 – Psoriasis; ICD-10-CM: T20–T25 – Burns and corrosions of external body surface, specified by site; CPT: 1014622, 44950, 44960, 44970 – Appendectomy; SNOMED: 80146002 – Appendectomy; ICD-10-CM: H91.1 – Presbycusis; RxNorm: 1514 – Betamethasone (topical product) | Any instance of Group 1B occurred on or before the first instance of Group 1A |
|  | Group 2A | Control index event meeting the two-encounter sensitivity definition | ICD-10-CM: Z00 – Encounter for general examination without complaint, suspected or reported diagnosis | Any instance of Group 2B occurred within 1 month on or after the first instance of Group 2A |
|  | Group 2B | Confirming healthcare encounter | Visit | Must have occurred within 1 month on or after the first instance of Group 2A |

| **Cohort** | **Query group** | **Definition** | **Codes** | **Temporal rule** |
| --- | --- | --- | --- | --- |
| **Atopic dermatitis (cases)** | Group 1A | Index event: first recorded diagnosis of atopic dermatitis | ICD-10-CM: L20 – Atopic dermatitis | Event between 1 Jan 2016 and 31 Dec 2018 |
|  | Group 1B | Exclusion criteria prior to cohort entry | ICD-10-CM: T20–T25 – Burns and corrosions of external body surface, specified by site; CPT: 1014622, 44950, 44960, 44970 – Appendectomy; SNOMED: 80146002 – Appendectomy; ICD-10-CM: H91.1 – Presbycusis; RxNorm: 1514 – Betamethasone (topical product); ICD-10-CM: L40–L45 – Papulosquamous disorders; ICD-10-CM: L49–L54 – Urticaria and erythema; ICD-10-CM: L21 – Seborrheic dermatitis; ICD-10-CM: L22 – Diaper dermatitis; ICD-10-CM: L23 – Allergic contact dermatitis; ICD-10-CM: L24 – Irritant contact dermatitis; ICD-10-CM: L25 – Unspecified contact dermatitis; ICD-10-CM: L26 – Exfoliative dermatitis; ICD-10-CM: L27 – Dermatitis due to substances taken internally; ICD-10-CM: L28 – Lichen simplex chronicus and prurigo; ICD-10-CM: L29 – Pruritus; ICD-10-CM: L30 – Other and unspecified dermatitis; ICD-9-CM: 690 – Erythematosquamous dermatosis; ICD-9-CM: 690.1 – Seborrheic dermatitis | Any instance of Group 1B occurred on or before the first instance of Group 1A |
| **Non-atopic dermatitis controls** | Group 1A | Index event: encounter for general examination without complaint, suspected or reported diagnosis | ICD-10-CM: Z00 – Encounter for general examination without complaint, suspected or reported diagnosis | Event between 1 Jan 2016 and 31 Dec 2018 |
|  | Group 1B | Exclusion criteria prior to cohort entry | ICD-10-CM: T20–T25 – Burns and corrosions of external body surface, specified by site; CPT: 1014622, 44950, 44960, 44970 – Appendectomy; SNOMED: 80146002 – Appendectomy; ICD-10-CM: H91.1 – Presbycusis; RxNorm: 1514 – Betamethasone (topical product); ICD-10-CM: L40–L45 – Papulosquamous disorders; ICD-10-CM: L49–L54 – Urticaria and erythema; ICD-10-CM: L21 – Seborrheic dermatitis; ICD-10-CM: L22 – Diaper dermatitis; ICD-10-CM: L23 – Allergic contact dermatitis; ICD-10-CM: L24 – Irritant contact dermatitis; ICD-10-CM: L25 – Unspecified contact dermatitis; ICD-10-CM: L26 – Exfoliative dermatitis; ICD-10-CM: L27 – Dermatitis due to substances taken internally; ICD-10-CM: L28 – Lichen simplex chronicus and prurigo; ICD-10-CM: L29 – Pruritus; ICD-10-CM: L30 – Other and unspecified dermatitis; ICD-9-CM: 690 – Erythematosquamous dermatosis; ICD-9-CM: 690.1 – Seborrheic dermatitis; ICD-10-CM: L20 – Atopic dermatitis | Any instance of Group 1B occurred on or before the first instance of Group 1A |
| **S6** | | | | |
| **Cohort** | **Query group** | **Definition** | **Codes** | **Temporal rule** |
| **Atopic dermatitis (cases)** | Group 1A | Index event: first recorded diagnosis of atopic dermatitis | ICD-10-CM: L20 – Atopic dermatitis | Event between 1 Jan 2016 and 31 Dec 2018 |
|  | Group 1B | Exclusion criteria prior to cohort entry | ICD-10-CM: L40 – Psoriasis; ICD-10-CM: T20–T25 – Burns and corrosions of external body surface, specified by site; CPT: 1014622, 44950, 44960, 44970 – Appendectomy; SNOMED: 80146002 – Appendectomy; ICD-10-CM: H91.1 – Presbycusis; RxNorm: 1514 – Betamethasone (topical product) | Any instance of Group 1B occurred on or before the first instance of Group 1A |
| **Non-atopic dermatitis controls** | Group 1A | Index event: first recorded diagnosis of acne vulgaris | ICD-10-CM: L70.0 – Acne vulgaris | Event between 1 Jan 2016 and 31 Dec 2018 |
|  | Group 1B | Exclusion criteria prior to cohort entry | ICD-10-CM: L40 – Psoriasis; ICD-10-CM: T20–T25 – Burns and corrosions of external body surface, specified by site; CPT: 1014622, 44950, 44960, 44970 – Appendectomy; SNOMED: 80146002 – Appendectomy; ICD-10-CM: H91.1 – Presbycusis; RxNorm: 1514 – Betamethasone (topical product) | Any instance of Group 1B occurred on or before the first instance of Group 1A |

| **S7** | | | | |
| --- | --- | --- | --- | --- |
| **Cohort** | **Query group** | **Definition** | **Codes** | **Temporal rule** |
| **Atopic dermatitis (cases)** | Group 1A | Index event: first recorded diagnosis of atopic dermatitis | ICD-10-CM: L20 – Atopic dermatitis, aged <18 years | Event between 1 Jan 2016 and 31 Dec 2018 |
|  | Group 1B | Exclusion criteria prior to cohort entry | ICD-10-CM: L40 – Psoriasis; ICD-10-CM: T20–T25 – Burns and corrosions of external body surface, specified by site; CPT: 1014622, 44950, 44960, 44970 – Appendectomy; SNOMED: 80146002 – Appendectomy; ICD-10-CM: H91.1 – Presbycusis; RxNorm: 1514 – Betamethasone (topical product) | Any instance of Group 1B occurred on or before the first instance of Group 1A |
| **Non-atopic dermatitis controls** | Group 1A | Index event: encounter for general examination without complaint, suspected or reported diagnosis | ICD-10-CM: Z00 – Encounter for general examination without complaint, suspected or reported diagnosis, aged <18 years | Event between 1 Jan 2016 and 31 Dec 2018 |
|  | Group 1B | Exclusion criteria prior to cohort entry | ICD-10-CM: L40 – Psoriasis; ICD-10-CM: T20–T25 – Burns and corrosions of external body surface, specified by site; CPT: 1014622, 44950, 44960, 44970 – Appendectomy; SNOMED: 80146002 – Appendectomy; ICD-10-CM: H91.1 – Presbycusis; RxNorm: 1514 – Betamethasone (topical product) | Any instance of Group 1B occurred on or before the first instance of Group 1A |

| **S8** | | | | |
| --- | --- | --- | --- | --- |
| **Cohort** | **Query group** | **Definition** | **Codes** | **Temporal rule** |
| **Atopic dermatitis (cases)** | Group 1A | Index event: first recorded diagnosis of atopic dermatitis | ICD-10-CM: L20 – Atopic dermatitis, aged ≥18 years | Event between 1 Jan 2016 and 31 Dec 2018 |
|  | Group 1B | Exclusion criteria prior to cohort entry | ICD-10-CM: L40 – Psoriasis; ICD-10-CM: T20–T25 – Burns and corrosions of external body surface, specified by site; CPT: 1014622, 44950, 44960, 44970 – Appendectomy; SNOMED: 80146002 – Appendectomy; ICD-10-CM: H91.1 – Presbycusis; RxNorm: 1514 – Betamethasone (topical product) | Any instance of Group 1B occurred on or before the first instance of Group 1A |
| **Non-atopic dermatitis controls** | Group 1A | Index event: encounter for general examination without complaint, suspected or reported diagnosis | ICD-10-CM: Z00 – Encounter for general examination without complaint, suspected or reported diagnosis, aged ≥18 years | Event between 1 Jan 2016 and 31 Dec 2018 |
|  | Group 1B | Exclusion criteria prior to cohort entry | ICD-10-CM: L40 – Psoriasis; ICD-10-CM: T20–T25 – Burns and corrosions of external body surface, specified by site; CPT: 1014622, 44950, 44960, 44970 – Appendectomy; SNOMED: 80146002 – Appendectomy; ICD-10-CM: H91.1 – Presbycusis; RxNorm: 1514 – Betamethasone (topical product) | Any instance of Group 1B occurred on or before the first instance of Group 1A |
