## Supplementary material for "Incident psoriasis in atopic dermatitis: A large-scale cohort study of disease- and treatment-associated risks": Sup Tab 2

**Supplement Table 2.** Cohort construction, index event definitions, and temporal rules for the primary and sensitivity analyses for Aim 2.

| **Primary analysis** | | | | |
| --- | --- | --- | --- | --- |
| **Cohort** | **Query group** | **Definition** | **Codes** | **Temporal rule** |
| **Biologic treatment cohort (cases)** | Group 1A | Index treatment: first recorded exposure to an AD-targeted biologic | RxNorm: 2589225 – Tralokinumab; 1876376 – Dupilumab; 2693758 – Lebrikizumab | Exposure on or after 1 Jan 2017 |
|  | Group 1B | Exclusion of alternative systemic treatments prior to cohort entry | RxNorm: 2591476 – Abrocitinib; 2196092 – Upadacitinib; 2047232 – Baricitinib; 6851 – Methotrexate; 3008 – Cyclosporine; 1256 – Azathioprine; 68149 – Mycophenolate mofetil | Any instance of Group 1B occurred on or before the first instance of Group 1A |
|  | Group 2A | Biologic exposure in patients with underlying atopic dermatitis | RxNorm: 1876376 – Dupilumab; 2693758 – Lebrikizumab; 2589225 – Tralokinumab | Any instance of Group 2B occurred within 1 month on or before the first instance of Group 2A |
|  | Group 2B | Atopic dermatitis diagnosis linked to treatment exposure | ICD-10-CM: L20 – Atopic dermatitis | Must have occurred within 1 month on or before the first instance of Group 2A |
|  | Group 3A | Incident atopic dermatitis anchor | ICD-10-CM: L20 – Atopic dermatitis | Incident AD definition |
|  | Group 3B | Exclusion criteria prior to incident AD | ICD-10-CM: T20–T25 – Burns and corrosions of external body surface, specified by site; ICD-10-CM: H91.1 – Presbycusis; ICD-10-CM: L40.0 – Psoriasis vulgaris; CPT: 1014622, 44950, 44960, 44970 – Appendectomy; SNOMED: 80146002 – Appendectomy | Any instance of Group 3B occurred on or before the first instance of Group 3A |
| **Conventional systemic immunosuppressant treatment cohort (controls)** | Group 1A | Index treatment: first recorded exposure to a conventional systemic immunosuppressant | RxNorm: 6851 – Methotrexate; 1256 – Azathioprine; 68149 – Mycophenolate mofetil; 3008 – Cyclosporine | Exposure on or after 1 Jan 2017 |
|  | Group 1B | Exclusion of biologic or JAK inhibitor exposure prior to cohort entry | RxNorm: 2693758 – Lebrikizumab; 1876376 – Dupilumab; 2591476 – Abrocitinib; 2589225 – Tralokinumab; 2196092 – Upadacitinib; 2047232 – Baricitinib | Any instance of Group 1B occurred on or before the first instance of Group 1A |
|  | Group 2A | Conventional systemic immunosuppressant exposure in patients with underlying atopic dermatitis | RxNorm: 6851 – Methotrexate; 1256 – Azathioprine; 68149 – Mycophenolate mofetil; 3008 – Cyclosporine | Any instance of Group 2B occurred within 1 month on or before the first instance of Group 2A |
|  | Group 2B | Atopic dermatitis diagnosis linked to treatment exposure | CD-10-CM: L20 – Atopic dermatitis | Must have occurred within 1 month on or before the first instance of Group 2A |
|  | Group 3A | Incident atopic dermatitis anchor | ICD-10-CM: L20 – Atopic dermatitis | Incident AD definition |
|  | Group 3B | Exclusion criteria prior to incident AD | ICD-10-CM: T20–T25 – Burns and corrosions of external body surface, specified by site; ICD-10-CM: H91.1 – Presbycusis; ICD-10-CM: L40.0 – Psoriasis vulgaris; CPT: 1014622, 44950, 44960, 44970 – Appendectomy; SNOMED: 80146002 – Appendectomy | Any instance of Group 3B occurred on or before the first instance of Group 3A |

The cohort construction logic for Aim 2 followed that of Aim 1, with treatment-specific adaptations outlined here.
